# Red cell folate status among a subset of Canadian children with Down Syndrome post-fortification

**DOI:** 10.1101/2021.10.06.21264479

**Authors:** Joan Jory

## Abstract

**Background:** Trisomy 21 or Down Syndrome (DS) is associated with altered methylation pathways. Children with DS may therefore represent a population subgroup with vulnerability to increased exposures to folic acid, which is involved in one-carbon metabolism. Folic acid (FA) fortification of flour and maternal FA supplementation are intended to reduce neural tube defects related to folate deficiency. The interventions have been widely successful in Canada. Emerging evidence suggests that higher FA exposures may also have potential negative consequences, including implications for DNA methylation. This retrospective chart review provides insight on the RBC folate status of a subset of Canadian children and infants with DS, post-fortification.

**Method:** Children with DS in 2 Canadian provinces were assessed in the community. Access to RBC folate testing was variable, limiting sample size to 39 (n=27 for children ≤ 6 years; n=12 for children 6-18 years). All children with DS and an RBC folate result were included. Use of FA-containing supplements and formula was documented.

**Results:** Among children 6-18 years, 100% had RBC folates > 1000 nmol/L, 50% were > 2000 nmol/L and 25% had levels above the reporting limit. Among the younger children (< 6 years), 52 % had RBC folates >2000 nmol and 2 children exceeded 3000 nmol/L. Among exclusively breast-fed infants (<12 months), 100% had RBC folates > 1000 nmol/L and 50% had levels > 2000 nmol/L, suggestive of in-utero or maternal exposures. RBC folate status among this subset of Canadian children with DS is higher than documented for the larger Canadian population, and higher than among US children with DS.

**Conclusions:** Young Canadian children with DS demonstrated high post-fortification RBC folate status. RBC folate status was higher than reported for the larger Canadian population, and higher than for US children with Down Syndrome. Consumption of folic acid-containing formula and/or supplements was relatively low among these Canadian children with DS, suggesting maternal FA supplements and/or FA-fortified foods may be important etiological factors. A larger, prospective study is needed to validate these results, and to explore potential health implications among this vulnerable population.

## INTRODUCTION

Mandatory folic acid fortification of flour was introduced to Canada in 1998 with a primary aim of reducing neural tube defects. The subsequent 46% reduction in overall neural tube defects is an important public policy success [17]. Folic acid fortification of flour was also accompanied by increased intakes of folic acid from multivitamins and maternal folic acid supplements in Canada [12]. The combined effects of increased folate-rich fruit and vegetable intakes alongside folic acid-fortified functional foods, together increased maternal folic acid supplementation during pregnancy and two decades of mandatory folic acid fortification of flour, have contributed to high red blood cell (RBC) folate levels among a larger-than-expected percentage of the Canadian population aged 6 to 79 years [1, 13].

Children with Down Syndrome may represent a group particularly vulnerable to excess folic acid exposures. Gene-dosing effects of the trisomy may alter micronutrient use related to production of enzymes coded on chromosome 21. Some of these enzymes, including cystathione beta synthase [36] and glutathione peroxidase [9], are involved in critical folate pathways [15]. Thus, higher exposures to folic acid may have unintended consequences among children with Down Syndrome. These may include the masking of occult B12 deficiency [7, 8] and alteration of DNA methylation, potentially impacting both pediatric neurodevelopment and epigenetic programming of premature aging [4, 29, 30].

The current study is a retrospective chart review of a subset of children with Down Syndrome from diverse communities in 2 Canadian provinces, initiated after a trend of high RBC folates was observed during community nutrition assessment.

## MATERIALS AND METHODS

### Patient inclusion and characteristics

The children with Down Syndrome were under medical supervision through physicians across communities in Ontario and New Brunswick, representing a diverse patient population. RBC folate tests were ordered by physicians as part of a screening panel for nutrition assessment at a community clinic in Guelph, Ontario. All children (0-18 years) with Down Syndrome and a baseline RBC folate result seen at the clinic between 2003 and 2016 were included; no child with Down Syndrome and an RBC folate result was excluded. Serum B12 was not routinely assessed.

Overall, 39 children met the inclusion criteria; 21 were male and 18 were female. There were two primary age groups of 0-5.9 years (n=27) and 6-18 years (n=12).

### RBC folate testing sites and interpretive reference ranges

RBC folate analyses were carried out in community and hospital labs across Ontario, and in New Brunswick. Interpretive reference ranges and sufficiency cut-offs for RBC folate results varied among the labs, and are documented by site and year. Assay and machine type are also documented.

### Supplement intake history

Consumption history of folic acid-containing supplements, oral formula and/or tube feeds was solicited for all children at intake. Differentiation between Down Syndrome-specific and general supplements was made. This information was numerically coded: 0=no supplements or formulas, 1=oral formula feeding, 2=G-tube formula feeding, 3=Down Syndrome-specific supplement (containing methylation co-factors), 4=other folic acid-containing vitamins. Retrospective data on specific folic acid dose and frequency of intake was not available. Parent perception of recall accuracy for supplement use declined with increasing age of the child at intake.

Use of maternal prenatal folic acid-containing supplements was also solicited. Maternal ability to retrospectively recall prenatal supplement use at intake was limited, and insufficient data was available for inclusion in this retrospective review.

### Statistical analysis

Group differences in RBC folate status by age (<6 vs 6-18 years), gender and supplemental folic acid exposure were assessed using the student T-Test. Linear correlations between RBC folate and continuous variables were assessed via the Pearson Correlation equation.

## RESULTS

### RBC folate results

The RBC folate results and additional characteristics for all children are presented in Table 1, in order of age at time of RBC folate testing. Ninety two percent (92%) of all children had RBC folate levels exceeding 1000 nmol/L. Fifty one percent (51%) had levels exceeding 2000nmol/L.

**Table 1:**
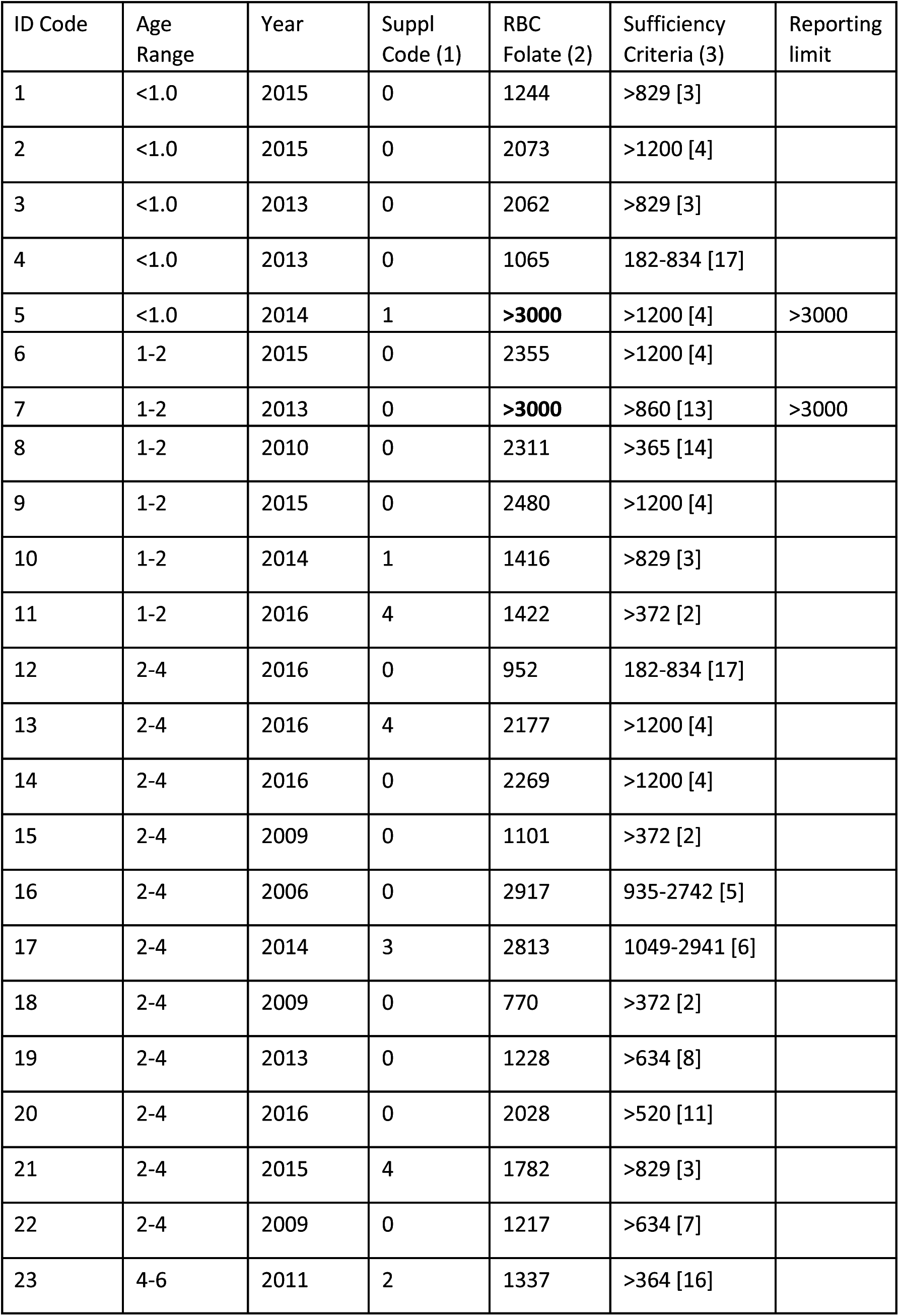
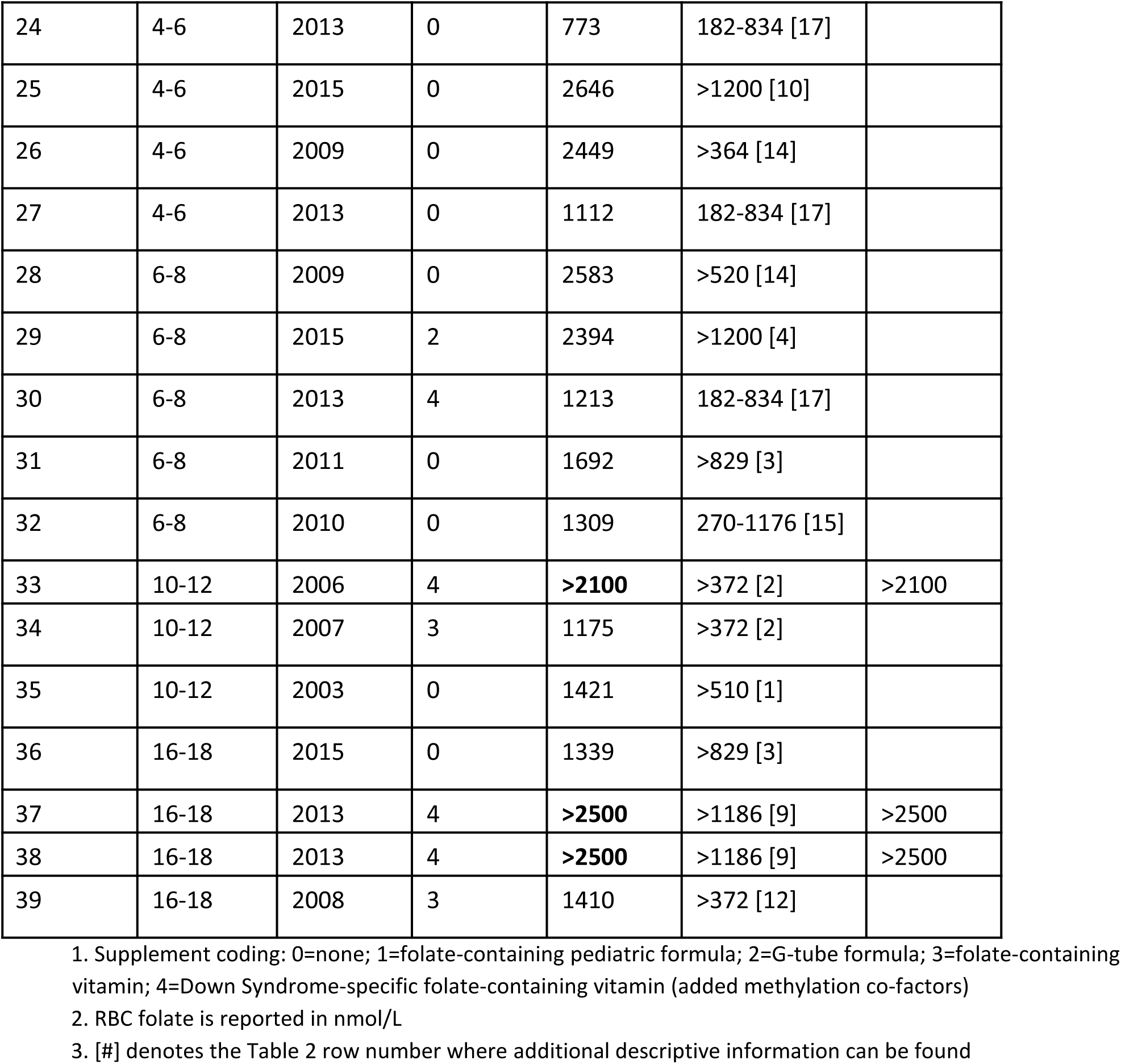
RBC Folate results

Some labs had an upper reporting limit for RBC folate outcomes. Five children (13%) had RBC folates above the upper reporting limit of the respective labs: two children exceeded an upper reporting limit of > 3000 nmol/L; 1 and 2 children exceeded the upper reporting limits of > 2100 and > 2500 respectively. During statistical analyses, these upper reporting limits were artificially substituted as the respective child’s RBC folate level. Their true RBC folate level was unknown but likely higher.

There were no significant gender differences in RBC folate status. There were no significant differences in RBC folate level between children under than 6 years and children 6-18 years. Neither child age nor year of RBC folate testing were significant explanatory correlates for RBC folate status.

### Folic Acid exposures

There was no recent exposure to folic acid-containing formulas or multivitamins at the time of folate testing among 74% of the younger children (< 6 years). Among the older children (6-18 years), 42 % had no supplemental folic acid exposure at time of testing. Overall, children consuming folic acid-containing supplements or formulas had higher RBC folate status (2100 +/- 555 nmol/L) compared to children with no supplemental folic acid exposures (1661 +/- 680 nmol/L; p<0.03). Specialized Down Syndrome formulas containing methylation co-factors were in use among 18% of the children overall.

### Laboratory reference ranges for RBC folate

The folate sufficiency cut-offs, or reference ranges where employed, for the respective RBC folate testing sites are presented in Table 2 by lab site, machine used and year of testing. Assay type is also noted. Some labs documented a folate sufficiency cut-off only. Some testing sites altered their reference ranges between 2003 and 2016; these changes are documented. Some labs changed ownership and name over time; the name documented in Table 2 is that provided by the lab at the time of testing.

**Table 2:** RBC sufficiency diagnostics by lab, machine type, and year.

| Laboratory | Machine + Method ( ) | Year | Sufficiency Cutoff or Range |
| --- | --- | --- | --- |
| 1.CML Labs | (4) Bio-Rad Quantaphase RIA | 2003 | >510 |
| 2.CML Labs | (4) Bio-Rad Quantaphase RIA | 2006-2009 | >372 |
| 3.CML Labs | (3) Beckman Coulter | 2013-2015 | >829 |
| 4.Gamma-Dynacare | (5) Roche Elecsys | 2014-2016 | >1200 |
| 5.Grand River Hospital | (5) Roche Elecsys | 2006 | 935-2742 |
| 6.Life Labs | (4) Bio-Rad Quantaphase RIA | 2009 | >634 |
| 7.Life Labs | (3) Beckman Coulter | 2013 | >634 |
| 8.Life Labs | (3) Beckman Coulter | 2013 | >1186 |
| 9.Life Labs | (3) Beckman Coulter | 2015 | >1200 |
| 10.London Health Sciences Corp | (5) Roche Elecsys | 2014 | 1049-2941 |
| 11.McMaster Hospital | (1) Abbott Architect | 2008-2009 | >520 |
| 12.MDS | (4) Bio-Rad Quantaphase RIA | 2008 | >372 |
| 13.Quest Diagnostic | (2) Advia Centaur | 2014 | >860 |
| 14.Saint John Regional Hospital | (5) Roche Elecsys | 2009-2010 | >365 |
| 15.Toronto Hospital for Sick Children | (2) Advia Centaur | 2010 | 270-1176 |
| 16.Toronto Hospital for Sick Children | (2) Advia Centaur | 2011 | >364 |
| 17.Toronto Hospital for Sick Children | (1) Abbott Architect | 2013,<br>2016 | 182-834 |

- (1) - Abbott Architect: Chemiluminescent microparticle immunoassay - (2) - Advia (Bayer) Centaur: Chemiluminescent competitive immunoassay (CLIA) - (3) - Beckman Coulter (Canada): Chemiluminescent immunoassay (CLIA) - (4) - Bio Rad: Quantaphase II Radioimmunoassay (RIA) - (5) - Roche Elecsys: Electro-chemiluminescence immunoassay

Overall, the lab reference ranges demonstrate rising cut-off criteria for folate sufficiency between 2003 and 2016.

## DISCUSSION

### Diagnostic Laboratory Reference Ranges

Red blood cell folate is a relatively stable biomarker of body folate status, useful for monitoring response to intervention strategies [23]. The current study identifies wide variations in interpretive criteria for RBC folate assessment at community and hospital labs in different regions of central and eastern Canada. All labs employed some form of immuno-assay; none were using a microbiological assay method at the time of testing.

Folate deficiency cut-offs between sites varied almost 7-fold, from as low as 182 nmol/L (Toronto Hospital for Sick Children, 2013) to 1200 nmol/L (Life Labs, 2015.) Some labs included an upper interpretive cut-off for RBC folate. These also varied widely from 834 and 1176 nmol/L at Toronto’s Hospital for Sick Children to 2742 nmol/L (Grand River Hospital, 2006) and 2941 nmol/L (London Health Sciences, 2014). The ability to interpret folate implications will be impacted by variations in diagnostic methodology and reference criteria for determination of abnormal RBC folate status.

The high RBC folate levels among this group of Canadian children with Down Syndrome supports the need for ongoing monitoring of the RBC folate status of vulnerable population groups such as those with Down Syndrome or pregnant women. Several provinces (New Brunswick and British Columbia) have delisted RBC folate as a government-funded test. Reductions in provincially funded RBC folate testing are also occurring in Ontario [27].

### Comparison of Canadian Down Syndrome folate status with the larger Canadian population

In 2011, Colapinto et al published findings on the folate status of representative Canadians aged 6-79 years based on the 2007-2009 Canadian Health Measures survey (CMHS) [13]. They employed a cut-off of 305 nmol/L for folate deficiency, based on the 5^th^ percentile from the United States National Health and Nutrition Examination Survey (NHANES) of 1999-2004. They also established an upper folate cut-off of 1360 nmol/L, based on the 97^th^ percentile from the United States National Health and Nutrition Examination Survey of 1999-200.Using this range (305-1360 nmol/L), fewer than 1% of participating Canadians demonstrated folate deficiency. By contrast, 40% of participants had high RBC folate levels (>1360 nmol/L). In a 2015 update, the Colapinto team revisited the folate status of Canadians [12]. Using modified cut-off criteria representing the 90^th^, 95^th^ and 97^th^ percentile from the post-fortification (2007-2010) NHANES dataset, they found that 16% of Canadians 6-79 years old had RBC folate results above 1450 nmol/L, 6% were above 1800 nmol/L and 2% had RBC folate results above 2150 nmol/L.

In the current review, high folate status was more common among the children aged 6-18 years than was documented for the larger Canadian population [12, 13]. Specifically, 67% had RBC folates above the 1360 nmol/L cut-off used by the Colapinto team in 2011 [13]. Further, 42% exceeded the 2015 high folate cut-off of 2150 nmol/l, compared to 2% in the general Canadian population [12].

Among these older children (6-18 years), 58% were consuming folic acid-containing supplements or formulas at the time of testing. Eighty percent (80%) of children 6-18 years with RBC folates exceeding 2000 nmol/L had recent supplemental folic acid exposures. Thus, among this older age group, recent supplemental folic acid exposures may contribute to their higher RBC folate status.

By contrast, only 26% of the younger children (< 6 years) consumed folic acid-containing formula or supplements at time of testing. Despite the lower supplemental folic acid exposures among these younger children, RBC folates also exceeded published cut-offs suggesting indirect exposures through maternal foods and supplements or possibly folic acid-fortified baby cereals. Specifically, 89% had RBC folates above 1000 nmol/L and 63% had RBC folates greater than 1360 nmol/L [13]. Fifty-five percent (55%) exceeded the Colapinto team’s 2015 cut-off of 1450 nmol/L, 52% exceeded the 1800 nmol/L cut-off and 41% exceeded the highest cut-off of 2150 nmol/L [12].

Of particular concern is a subgroup of infants (<1 year; n=8) with Down Syndrome. Seventy-five percent (75%) had RBC folates above 2000 nmol/L, and 25% had RBC folates exceeding 3000 nmol/L. Only 1 child was consuming a folic acid-containing formula. The remaining 88% of infants were exclusively breast-fed and received no formula or vitamin preparations, suggesting in-utero folic acid exposure and/or maternal post-partum intake as etiological vectors.

The lowest RBC folate among these Canadian children with Down Syndrome (all ages combined) was 770 nmol/L, more than twice the 305 nmol/L cut-off used by for folate deficiency [13]. None met the Colapinto criteria for folate deficiency (<305 nmol/L) [13], and no child with Down Syndrome had an RBC folate level below the respective lab-specific cut-off for folate sufficiency (Table 1). Among 49%, RBC folates exceeded the CMHS 97^th^ percentile for Canadians aged 6-11 (1984 nmol/L) and 12-19 (1977 nmol/L) years respectively [12].

### Comparison of folate status among other children with Down Syndrome

There are few published data on RBC folate status among children with Down Syndrome. However, Funk et al [20] identified lower than expected RBC folates among 15 US children with Down Syndrome (median age 4.5 years) enrolled in a juvenile arthritis study. Mean RBC folate was lower among the Down Syndrome children, at 845.5 ± 403.5 nmol/L, compared to 1121.6 ± 512.3 nmol/L for the control group of neurotypical arthritis sufferers. By contrast, the mean RBC folate 1852 ± 724 nmol/L for the younger Canadian children (< 6 years) with Down Syndrome was more than 2-fold higher than that of the US children with Down Syndrome. A follow-up Funk publication on 85 children with Down Syndrome (median age 5 years) confirmed these findings of lower RBC folates compared to controls [19]. The US Down Syndrome children with folic acid-containing supplement exposures trended toward higher RBC folates than non-supplement users (p=0.07) [20], in a manner similar to that among Canadian children with Down Syndrome (p<0.03).

### Implications of high folate status among Canadian children with Down Syndrome

The presence of high RBC folates among young Canadian children with Down Syndrome may be of concern for several reasons. First, it suggests that cumulative increases in maternal folic acid (PteGlu) intakes from fortified foods and supplements may exceed the capacity to metabolize folic acid by the mother, the fetus or the young child. Pfeiffer et al found detectable levels of unmetabolized folic acid (UMFA) in >95% of Americans ≥ 1 year old from the 2007-2008 NHANE survey [37]. Higher levels (>1 nmol/L) were associated with smaller body size, and with total folic acid intake from diet and supplements [37]. By contrast UMFA was present in only 33% of US samples in the earlier 1999-2000 NHANES where, among vulnerable subgroups, it was associated with lower cognitive scores [34]. In Canada, UMFA has been detected in > 90% of maternal and umbilical cord blood samples [40].

Secondly, if folic acid is not well metabolized and excreted, additional prenatal supplementation among folate-replete populations may increase foetal UMFA and possibly contribute to unintended consequences. Wiens et al. identified a dose-dependent relationship of increasing UMFA with reduced neurite length and synapse formation during neurogenesis in a rat model of brain development [50]. De Crescenzo et al. have further identified decreased cerebral cortex thickness, decreased cortical projection neurons and delayed prenatal neurogenesis in the mouse model of folic acid excess [16]. Two research teams identified that high levels of folic acid supplementation can cause growth retardation and heart defects in a mouse model of embryonic development [32, 38]. Higher cord blood UMFA was associated with decreased fetal DNA hydroxymethylation among a cohort of pregnant Canadian women [39]. Choi et al. [11] urge that excess intake of unmetabolized folic acid may have important epigenetic effects, and that interactions between individual phenotype and excess UMFA may put specific groups at risk.

It is established that epimutations in folate, homocysteine and methylation pathways constitute an important risk factor for the development of Down Syndrome [5, 48]. Coppede et al. [14] confirmed that folate-related polymorphisms are also an important risk factor for Down Syndrome births among younger mothers (<35 years). It has also been demonstrated that supra-physiological folic acid can decrease intracellular levels of S-adenosylmethionine (SAM), a methyl donor, and alter DNA methylation in human cell lines [10]. Methylation pathways are altered in children with Down Syndrome; compared to their siblings, children with Down Syndrome may present with lower SAM and S-adenosyl-homocysteine (SAH) levels due to overexpression of the cystathionine B-synthase gene [41]. Unexpected hypermethylation of lymphocyte DNA may reflect increased methylation demands in Down Syndrome; in vitro addition of methylation cofactors (methionine, methylcobalamin, folinic acid, and di-methyl-glycine) to Down Syndrome lymphoblasts improved status of all metabolites along the methionine - homocysteine pathway [41]. Since folic acid metabolism requires methylation [47], high exposures to folic acid in children with Down Syndrome may critically increase competition for methyl groups along genetically compromised methylation pathways. High intakes of folic acid among mothers with undetected methylation-related polymorphisms may conceivably also influence risk of Down Syndrome pregnancies [26, 28].

There are also important interrelationships between folate and cobalamin (B12), although the question of whether and how high folic intake might impact B12 pathways is a subject of ongoing scientific inquiry [31, 44]. Ray et al. identified that, among a select group of Canadians one year after mandatory folate fortification, RBC folate levels were higher than expected and higher than the respective serum B12 levels [43]. Plasma homocysteine levels proved non-diagnostic for B12 deficiency among the study’s folic acid-fortified population [43]. The authors noted that diagnostic challenges could therefore render occult B12 deficiency during high folate exposure an emerging public health concern [43]. Altered methylation pathways are often associated with lower homocysteine production in children with Down Syndrome [24, 41]. Therefore, homocysteine levels may also be non-diagnostic for B12 deficiency in children with Down Syndrome, possibly leading to increased risk of occult B12 deficiency.

The diagnostic reliability of methylmalonic acid (MMA), an alternate biomarker of B12 status, may also be questionable. Notably, B12 status was low in 17% or marginal in 38% of a cohort of Canadian women early in pregnancy, despite MMA elevations in fewer than 2% and an absence of homocysteine elevations [49]. Neither maternal serum B12 nor B12 intakes (diet and supplements) were statistically related to either maternal or cord blood MMA levels [49]. Maternal serum B12 status was also unrelated to neonatal MMA in an Indian cohort [18].

Undiagnosed B12 deficiency can be associated with a range of abnormalities [2, 22]. In the US National Health and Nutrition Examination Survey (NHANES) 1999-2002, increasing biochemical folate concentration in the presence of low B12 was associated with increasing dose-dependent impairment in cognitive function among older adults [46]. No such effects were identified in the pre-folate fortification NHANE survey of 1991-1994 [45]. Conceivably, then, increased RBC folate status in Down Syndrome may influence the integrity of adult cognitive function.

During pregnancy, an increasing ratio of folate to B12 altered foetal anthropometrics with dose-dependent decreases in birth weight, birth length, and head and chest circumference among Indian women receiving folate supplements [21]. There is also biochemical interplay between B12 and folate status in the etiology of neural tube defect risk [6]. In both folate-fortified [42] and folate-unfortified populations [33], the lowest quartile of B12 status was also associated with a 2 to 3-fold increase in neural tube defect risk.

In children, congenital and acquired B12 deficiency can be associated with inadequate myelination, seizures, hypotonia, lethargy and microcephaly [25]. Among Italian children with Down Syndrome specifically, serum B12 levels < 442 pg/mL (326 pmol/L) were associated with lower IQ scores [3]. Although rising homocysteine levels were also negatively associated with some cognitive measures, B12 status did not correlate with homocysteine levels. Folate status showed a modest negative association with homocysteine, but only after removal of statistical outliers. Folate fortification in Italy was voluntary at the time of the study. Therefore, it is possible that higher RBC folate status in countries with mandatory folate fortification, such as Canada, may negatively impact developmental outcomes among children with Down Syndrome who have marginal B12 status, irrespective of whether true deficiency criteria are met.

The relationships between folate, B12, one-carbon metabolites and health outcomes are complex and unclear. This is especially so for children with Down Syndrome among whom these pathways differ from the larger population. High intakes of folic acid may therefore have important impacts among this vulnerable group.

## CONCLUSIONS

This retrospective chart review was limited in scope, and its findings may not reflect other populations of children with Down Syndrome. The sample size was small, there was no control group of children without Down Syndrome, data on B12 and other one-carbon pathway metabolites was not available, and retrospective recall of folic acid-containing supplements was unreliable.

Nevertheless, this appears to be the first documentation of high folate levels among young Canadian children with Down Syndrome, a long-term folic acid-fortified country. RBC folates were higher among these Canadian children with Down Syndrome than reported for the general Canadian population (6-79 years). Canadian children with Down Syndrome also demonstrated an RBC folate status more than 2-fold higher than US children with Down Syndrome.

These findings will need to be replicated by others, ideally in a prospective case-control manner and including more one-carbon pathway metabolites with a larger sample size. Continued RBC folate monitoring of the larger population and of vulnerable subgroups among folate-fortified countries such as Canada may help to better elucidate the complex relationships between increasing folate exposures and health outcomes. Children with Down Syndrome and mothers who have altered folate-homocysteine-methylation pathways may be at particular risk of potential adverse effects.

## Data Availability

All data related to this retrospective chart review has been included in Tables 1 and 2.

## Acknowledgements

The author wishes to acknowledge the collaboration of the primary care physicians through whom red blood cell folate testing was carried out, and wishes to express gratitude to the participating children with Down Syndrome and their families.

## Authorship

Joan Jory RD PhD is responsible for the manuscript and submission, and accepts responsibility for the accuracy and scientific integrity of the paper.

## Institutional Review Board Statement

The study was conducted according to the guidelines of the Declaration of Helsinki, and approved by the Institutional Review Board of IRB Services Ontario (renamed Advarra - https://www.advarra.com/services/irb-services/canadian-review-services/). Protocol # Pro00020565; Approval January 31-2017.

## Informed Consent Statement

All patient data was strictly de-identified/anonymized, and written patient consent was waived in accordance with the Office of the Information and Privacy Commissioner of Ontario [35].

## Data Sharing

All data related to this retrospective chart review has been included in Tables 1 and 2.

